# A spatial AI-based biomarker with independent prognostic value in early low- and intermediate- risk breast cancer

**DOI:** 10.64898/2026.10.07.26364961

**Authors:** Constance Boissin, Yujie Xiang, Bojing Liu, Johan Vallon-Christersson, Espen Walker, Johan Hartman, Mattias Rantalainen

## Abstract

**Background:** Patients with ER-positive/HER2-negative early breast cancer of low or intermediate grade are generally considered to have a favourable prognosis, yet a number will recur. This risk is poorly captured by standard clinicopathological factors. AI-based medical devices for whole slide image (WSI) analyses used for risk-stratification often predict on slide-level thereby overlooking spatial localisation of the biomarkers. We hypothesise that the spatial distribution of a previously reported AI-based marker provides independent prognostic value beyond its global score.

**Patients and Methods:** This Swedish multicentre cohort study includes 6106 patients with primary invasive breast cancer: 3078 from the Stockholm region (training) and 3028 from Skåne (independent test). Tile-level high-grade morphology scores were generated with ViT_DG, a vision-transformer adaptation of DeepGrade, and the presence of clusters of high-risk tiles localised at the invasive tumour front was defined as a binary biomarker. Primary analyses were pre-specified in the ER-positive/HER2-negative/lymph-node-negative and Grade 1-2 subgroup. Association with breast-cancer recurrence-free interval (BCRFI) and distant metastasis were assessed using multivariable Cox proportional hazards models (HR).

**Results:** In the primary subgroup (n=1536 training; n=1403 test), tumour front clusters were present in 37% and 39% of patients. Cluster-positive status was associated with a 2.30-fold increased risk in the external test. Cluster-negative patients achieved a 10-year BCRFI of 95.3%, equivalent to Grade 1 patients and better than Grade 2 (92.4%), whereas cluster-positive patients (90.2%) did worse than the Grade 3 reference. The association was retained in patients classified as low-risk by ViT_DG (test set multivariable HR 2.86), identifying high-risk patients not captured by the global score.

**Conclusions:** The absence of high-risk morphology clusters at the tumour front is an independent prognostic indicator, identifying a low-risk group independently of the global AI biomarker classification. More broadly, preserving the spatial distribution of AI-derived predictions enables prognostic stratification beyond slide-level aggregation.

ClinicalTrials.gov identifier: NCT02306096

## Introduction

Breast cancer is the most commonly diagnosed cancer in women worldwide and a leading cause of cancer-related mortality^1^. Accurate patient stratification is key for clinical decision making, especially when determining the need for adjuvant chemotherapy. While high-risk patients, for example those with Nottingham Histological Grade (NHG) 3, or those with triple negative breast cancers, are clear candidates for aggressive treatment, other subgroups are more biologically heterogeneous, which subclassification requires special attention. This is particularly true for cases with oestrogen receptor-positive, human epidermal growth factor receptor 2-negative (ER+/HER2−) tumours, including NHG 2 cases. Patients with NHG1 tumours are typically considered low-risk and treated with endocrine treatment alone, without being candidates for any genomic profiling. Yet, some patients may present with some high-risk patterns that are currently not detected with standard assessments. NHG2 tumours are a very heterogeneous and intermediate group where treatment decisions are uncertain as highlighted by the 2023 St Gallen consensus, which shows that treatment recommendations for this group increasingly depend on the integration of tumour stage, histological grade, proliferative activity and genomic risk profiling^2^. Genomic assays such as Oncotype DX^3^ and Prosigna^4,5^ have been developed and are in clinical use in many contexts to improve risk stratification and enable the de-escalation of chemotherapy in lower-risk patients, however, they remain expensive and time-consuming and are based on bulk-average molecular profiles.

Digital pathology is now widely adopted in clinical context together with the use of high-resolution whole-slide histopathology images (WSIs), which enable AI and deep learning applications and analyses of WSIs^6^. The use of WSIs to risk-stratify early breast cancer patients has now been shown in a number of studies^7–10^ and further developed into commercial products (Ataraxis Breast, Spotlight Medical, ArteraAI Breast Cancer Test, Stratipath Breast). The previously reported DeepGrade model^9^ – a deep learning ensemble of convolutional neural networks trained to discriminate between NHG1 and NHG3 morphological patterns and thereafter subtype all patients into high- and low-risk groups, showed a performance comparable in effect size to established gene expression profiling assays. This model has been further developed into a regulatory-approved (CE-marked) AI-based medical device and has been further validated^11,12^. More recently, vision-transformers (ViT) models together with attention layers^13^ have improved the diagnostic performances at the patient level, however, they come with the key limitation that all tile-level predictions are aggregated into a single slide-level score through averaging, or attention-based multiple instance learning (ABMIL) frameworks^14^. These aggregation strategies deliberately overlook the spatial coordinates of individual tile predictions and dismiss spatial tumour architecture as a whole, losing information about where within the tumour a given morphological pattern is located.

Spatial intratumour heterogeneity (ITH) is the variation in cellular morphology, molecular phenotype and microenvironmental composition across different regions of the tumour. It is a well-established feature of breast cancer and a driver of disease progression and treatment resistance^15,16^. A specific example is the spatial localisation of TILs (tumour infiltrating lymphocytes) that carry prognostic value beyond the global density of the marker^17–19^. The invasive tumour front, or the outer region of the tumour where malignant cells interact with surrounding tissues, is the site of active stromal invasion, where epithelial-mesenchymal transition is enriched and cancer cells acquire migratory and invasive phenotypes^20,21^. Spatial zonation was demonstrated in breast cancer through spatial transcriptomics^22^, and tumour front characteristics such as tumour budding have been associated with prognosis^23,24^.

Despite the biological knowledge behind ITH affecting prognosis, few studies have focused on developing and validating spatially-resolved AI-based markers, with the potential to identify patients at increased risk of recurrence. In this study, we evaluate whether the spatial localisation of a previously defined AI-biomarker trained for high-grade morphology adds prognostic value independently of the biomarker itself, potentially contributing independent information that could assist in adjuvant treatment decisions, including decisions for treatment de-escalation which might be performed with greater confidence.

## Methods

### Study design and data

This is a retrospective analysis of two Swedish breast cancer cohorts. The study comprises female patients diagnosed with primary invasive breast cancer from five different Swedish hospitals sites across two regions. The training cohorts consisted of patients from the Stockholm region: Stockholm South General Hospital (SOS, 2012-2018, n=2902) and Karolinska Hospital, Solna (KS-Solna, 2009-2018, n=1849), for a total of n=4751 patients before exclusions. The external test cohort consisted of patients from three hospitals in the Skåne region (2010-2019): Lund (n=1235), Malmo (n=1650), and Kristianstad (n=1586), for a total of n=4471 before exclusions. These patients were all enrolled in the Sweden Cancerome Analysis Network – Breast (SCAN-B) study (ClinicalTrials.gov NCT02306096), a prospective, population-based, multicentre study enrolling patients with primary breast cancer in the Skåne region since 2010^25^. Only patients who had an available formalin-fixed paraffin-embedded (FFPE) haematoxylin and eosin (H&E)-stained slide from surgically resected tumours were included. Patients were excluded if no tumour was identified on the digitised WSI, if the WSI was of insufficient quality, if contained fewer than 100 tumour tiles after preprocessing, if the tumour front represented more than 85% of the tumour area, or if clinical variables required for multivariable analysis were missing. One slide was selected per patient based on availability, matched immuno-histochemistry slides, or largest calculated tumour size. An overview of the training and test cohorts is presented in Figure 1, while detailed presentations for each site are in Supplementary Figure S1. The main subgroup for analysis in this study is patients whose ER+/HER2-tumours are lymph node negative and of NHG 1 or 2, as it is those in whom indication for adjuvant chemotherapy cannot be decided from clinicopathological factors alone and therefore in whom gene expression assay are recommended according to the ESMO guidelines^26^.

**Figure 1.**
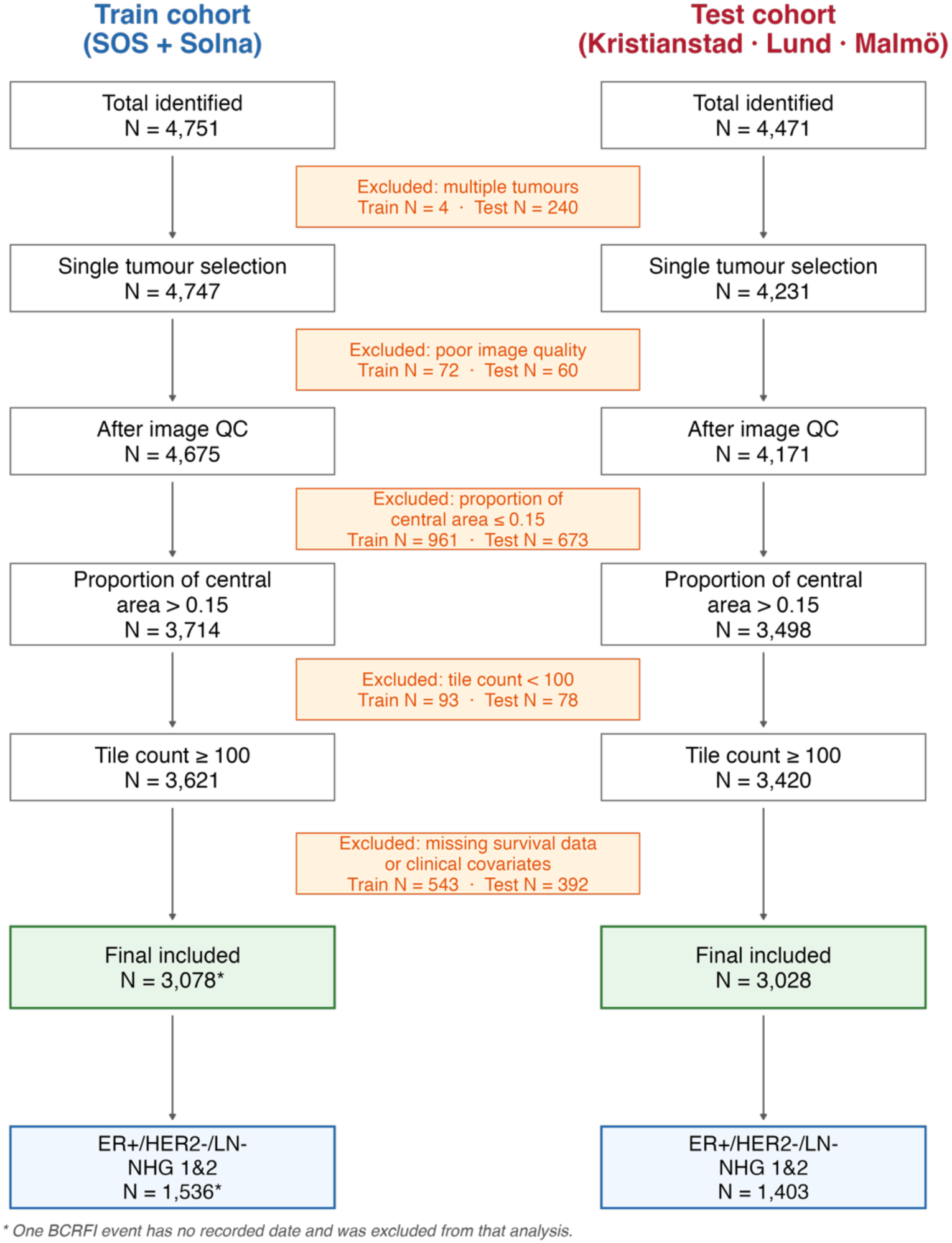
Flowchart illustrating the process of patient selection for the train and test splits.

### WSI preprocessing

Slides were digitised using Hamamatsu NanoZoomer XR, Hamamatsu NanoZoomer S360, Philips IntelliSite Pathology Solution 3.2 Ultra Fast Scanner, or Aperio GT450X at 40X magnification (0.226 µm/pixel), and then downsampled to an effective resolution of 20X (0.452 µm/pixel). Slides were then tiled into non-overlapping image patches (i.e. tiles) of 224X224 pixels, corresponding to a physical area of approximately 101µm X 101µm per tile. Tissue detection was performed by extracting lower-resolution representations of each WSI using OpenSlide^27^, transforming RGB to HSV colour space and applying Otsu’s thresholding to the saturation channel^28^. Tiles with less than 70% of tissue or a Laplacian variance smaller than 300 were excluded. Colour normalisation was performed at the WSI level using the Macenko method^29^, and previously described in^9^ to reduce systematic differences between scanners and sites. Tile-level feature embeddings of size 1024 were extracted using UNI, a self-supervised foundation model pre-trained on a large multi-cancer histopathology WSIs^13^.

Tumour regions within each WSI were defined using a tile-level binary classification model trained on UNI feature vectors, discriminating between invasive cancer regions and non-cancer tissue, developed using n=909 pathologist-annotated WSIs from the SOS cohort. Each segmented tumour region was subdivided into two anatomical zones: the tumour front (outermost 0.9mm from the tumour boundary, measured inward without tile overlap) and the tumour centre (all remaining tiles beyond the peripheral zone). The width of the tumour front zone (0.9 mm) was chosen to approximate one field of view at 20X.

### Spatial marker definition

Tile-level high-grade morphology scores were generated using ViT_DG, a re-optimised ViT adaptation of the DeepGrade model ^9^, using UNI-extracted feature vectors. A binary classification model was trained on NHG1 and NHG3 patients from SOS (n=922); with tile level scores representing similarity with high-grade morphology. ViT_DG obtained a tile-level AUC of 0.945 (training) and 0.875 (internal test), and slide-level AUC of 0.931 was obtained in the internal test set. Slide-level predictions were obtained using the 75th percentile of tile-level scores and the binary classification threshold was obtained using Youden’s J statistic^30^. The WEEP methodology (Wsi rEgion sElection aPproach)^31^ was applied to identify the minimal set of tiles driving the slide-level high-grade classification through iterative backward removal of highest-ranked tiles until the slide level score fell below the classification threshold. All tiles with a prediction score higher than the WEEP classification threshold were classified as high-risk tiles.

To identify whether the spatial representation of the AI-based biomarker was relevant in addition to the trained binary slide-based classification, we defined clusters of tiles that represent high-risk morphology and that could be present at the invasive tumour front. In order to identify those clusters, we first define the neighbouring areas of all tiles within the tumour front. We considered a neighbourhood as all tiles within four-tile steps allowing vertical, horizontal, or diagonal connectivity. A high-risk tumour front cluster (TFC) is defined if a high risk tile in the tumour front region has two thirds of its neighbouring tiles also classified as high risk. Patients were classified as cluster-positive if at least one such cluster was identified, and as cluster-negative otherwise. This binary variable constituted the primary spatial marker evaluated in the study. (See Figure 2).

**Figure 2.**
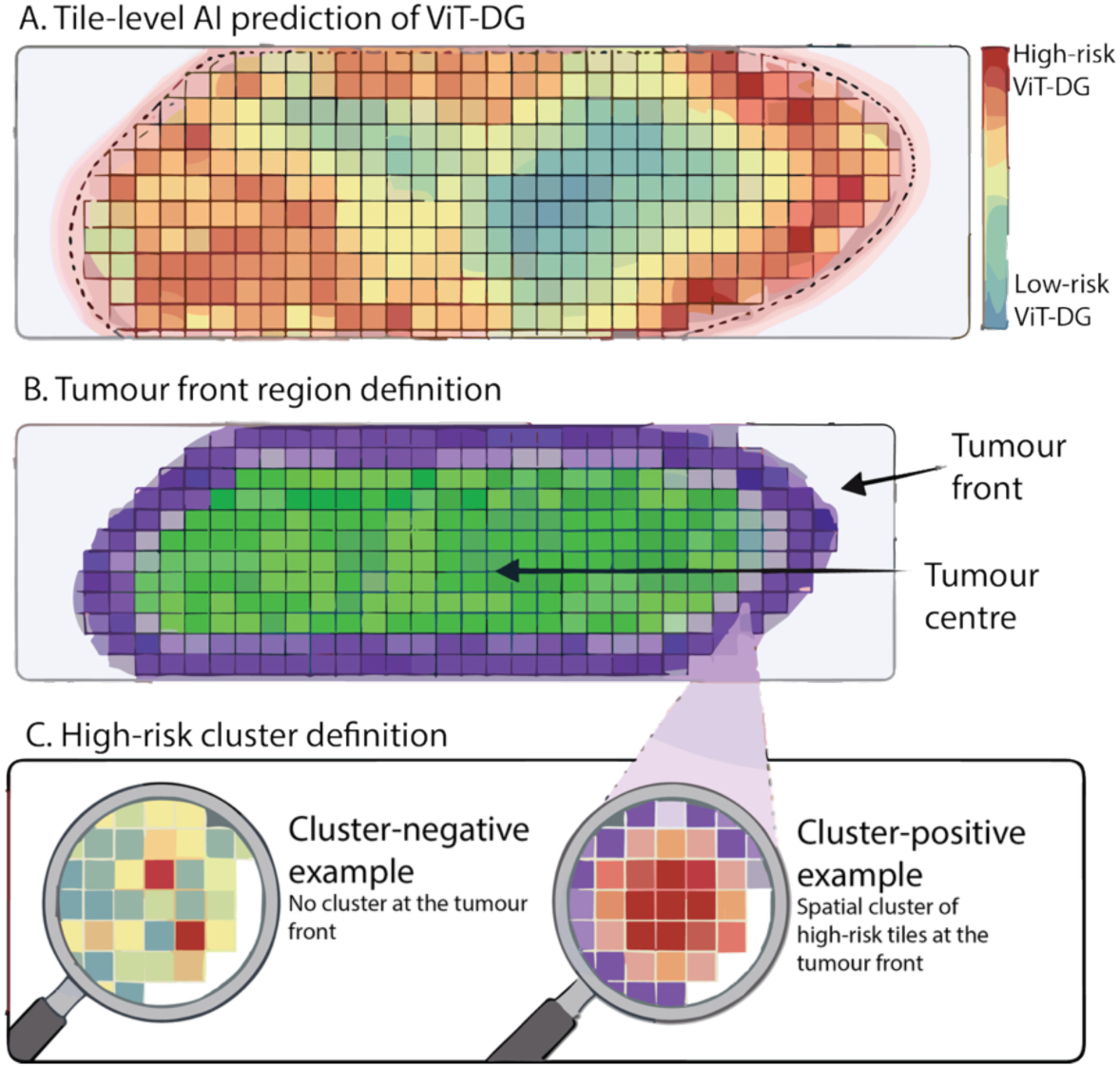
Schematic representation of the spatial tumour front clustering methodology. Overview of the spatial clustering approach used to identify the high-risk tumour front cluster positive patients. A) Tile-level risk predictions using ViT_DG for all tiles in a WSI as a continuous marker. WEEP backwards selection is then used to dichotomise tiles into high- and low-risks (not shown). B) The overall tumour region is defined by model prediction. It is then segmented into tumour centre and tumour front regions. C) High risk spatial clusters are identified in the tumour front area by detecting high-risk tiles that are surrounded by at least two thirds of neighbouring tiles which are also high risk.

The parameters defining the spatial marker were specified on biological and technical grounds rather than optimised against outcome. They were all defined in the training cohort (Stockholm), and never modified based on results from the test set.

### Statistical analyses

The primary outcome was breast cancer recurrence-free interval (BCRFI), defined as the time from initial diagnosis to the date of local, regional, or distant recurrence or contralateral breast cancer. A secondary outcome was distant-metastasis recurrence-free interval (DRFI), defined as the time from initial diagnosis to the date of distant metastasis. Kaplan–Meier curves were generated to visualise survival outcomes between cluster-positive and cluster-negative groups. Absolute survival differences in BCRFI and DRFI between cluster-positive and cluster-negative groups were estimated using the KM estimator log-rank tests at 5 and 10 years.

ER status was determined using immunohistochemical staining, with tumours classified as ER-positive if ≥10% of cells showed positive nuclear staining. HER2 status was determined by immunohistochemical staining and, where equivocal (IHC 2+), was confirmed by in situ hybridisation (ISH), according to local clinical guidelines. Tumours with IHC score 3+ or ISH amplification were classified as HER2-positive; all others as HER2-negative. Lymph node status was determined from pathological examination of the surgical specimen; patients were classified as lymph node-negative in the absence of nodal metastases.

Univariable and multivariable Cox proportional hazards regression models were used to estimate hazard ratios (HRs) with 95% confidence intervals (CIs) using time-since-cancer-diagnosis as underlying time scale. Covariates included in multivariable models for the primary subgroup analyses (ER+/HER2−/LN−/NHG 1–2) were age at diagnosis and tumour size (dichotomised as <20 mm versus ≥20 mm). Patients with missing data in any covariate were excluded from multivariable analyses.

Prognostic analyses were first performed in the full training cohort (Stockholm region) and subsequently validated in the external test set (Skåne region). The primary analyses were pre-specified for the subgroup of patients with ER-positive, HER2-negative, lymph-node negative, NHG 1 or 2 tumours (ER+/HER2−/LN−/NHG 1&2). Additional analyses were performed in the ViT_DG-low subgroup (ER+/HER2-/LN-patients classified as low-grade risk by ViT_DG) across the full unselected cohorts.

To compare the cluster-positive and cluster-negative groups, clinicopathological variables were compared using Fisher’s exact or chi-squared tests for categorical variables, and Student’s t-test or Mann–Whitney U test for continuous variables, as appropriate. Analyses were performed using R (version 4.5.2).

### Ethics approval and consent to participate

The SCAN-B study was approved by the Regional Ethical Review Board in Lund, Sweden (registration numbers 2009/658, 2010/383, 2012/58, 2013/459, 2014/521, 2015/277, 2016/541, 2016/742, 2016/944, 2018/267) and the Swedish Ethical Review Authority (registration numbers 2019-01252, 2024-02040-02). All patients in that study provided written informed consent prior to enrolment. All analyses were performed in accordance with patient consent and ethical regulations and decisions. The study and data from Stockholm were approved by the Swedish Ethical Review Authority (Etiksprovningsmyndigheten, Stockholm, Sweden, reference 2017/2106-31, with amendments 2018/1462-32 and 2019–02336); and no additional informed consent was required in accordance with ethical approval in this non-interventional collection and analysis of data from patient records. The study was performed in accordance with the Declaration of Helsinki.

## Results

### Patient cohorts

The training cohort comprised 3078 patients with primary invasive breast cancer from the two hospitals in the Stockholm region (Solna, N = 1471; Stockholm South General Hospital, N = 1607). The external test cohort comprised 3028 patients from three hospitals in the Skåne region (Kristianstad, N = 1198; Lund, N = 931; Malmo, N = 899). Clinicopathological characteristics of both cohorts are summarised in Supplementary Table S1. Patients were excluded according to the methods(site-specific flowchart in Supplementary Figure S1). Analyses focused on the pre-specified primary subgroup of patients with ER-positive, HER2-negative, lymph node-negative, NHG 1 or 2 tumours (ER+/HER2−/LN−/NHG 1&2), yielding n=1536 patients in the training cohort and n=1403 patients in the external test cohort, presented in Supplementary Table S2.

### Tumour front cluster prevalence

Within the training cohort, high-risk tumour front clusters were defined as neighbouring regions with at least two thirds of high-risk tiles within the invasive tumour front, were identified in 1769 of 3078 patients (57.5%) of the whole set and in 564 of 1536 patients (36.7%) of the primary (ER+/HER2−/LN−/NHG 1&2) subgroup. In the external test cohort, high risk clusters were identified in 1812 of 3028 patients (59.8%) of the whole set and 550 of 1403 patients (39.2%) of the primary subgroup. The remaining patients did not have any high-risk cluster detected at the tumour front and therefore were classified as cluster-negative. Tumour front cluster-positive patients showed higher proportions of ViT_DG-high tumours (73% versus 2%, p<0.001 in the combined train and test sets), higher Ki67 expression (median 20.0% versus 12.0%, p<0.001) a higher proportion of NHG2 tumours (88% versus 54%, p<0.001), and a larger tumour size (31% of tumours ≥20 mm in size versus 17% smaller, p < 0.001), whereas age and PR status were not significantly different between groups (Supplementary Figure S2).

### Prognostic value of tumour front clusters in the primary analysis subgroup

The prognostic performance of the tumour front clusters was identified in the training cohort (ER+/HER2-, LN-, NHG 1-2, N=1536; Supplementary Figure S3) with an absolute survival difference of 1.6% at 5 years and 3.6% at 10 years between the cluster-positive and the cluster negative subgroups (Table 1). This was then confirmed in the fully independent test cohort (ER+/HER2−, LN−, NHG 1–2 subgroup, N = 1403). Kaplan–Meier analysis confirmed a significantly worse BCRFI in cluster-positive patients (log-rank P < 0.001; Figure 3A), with an absolute difference of 3.2% at 5 years and a 5.1% difference at 10 years (Table 1). In univariable Cox regression, the presence of tumour front clusters was associated with a 2.34-fold increase in the training set and a 2.43-fold increase in the risk of recurrence in the test set (HR 2.43, 95% CI 1.52–3.89, P < 0.001), and cluster positive status remained a strong and statistically independent predictor in the multivariable analysis adjusting for age and tumour size (HR 2.30, 95% CI 1.43–3.70, p = 0.001; Figure 3B), with an effect size comparable to that observed in the training cohort (HR 1.81, 95% CI 1.01-3.25, P=0.048, Supplementary Figure S3A-B). The prognostic association was similarly confirmed for distant-metastasis recurrence-free interval (HR 3.17, 95% CI 1.53–6.53, p = 0.002 univariable; HR 2.85, 95% CI 1.37–5.92, p = 0.005 multivariable; Figure 3C-D), similar to what was seen in the training set (HR 2.04, 95% CI 1.02–4.10, p = 0.044; Supplementary Figure S3C-D).

**Figure 3.**
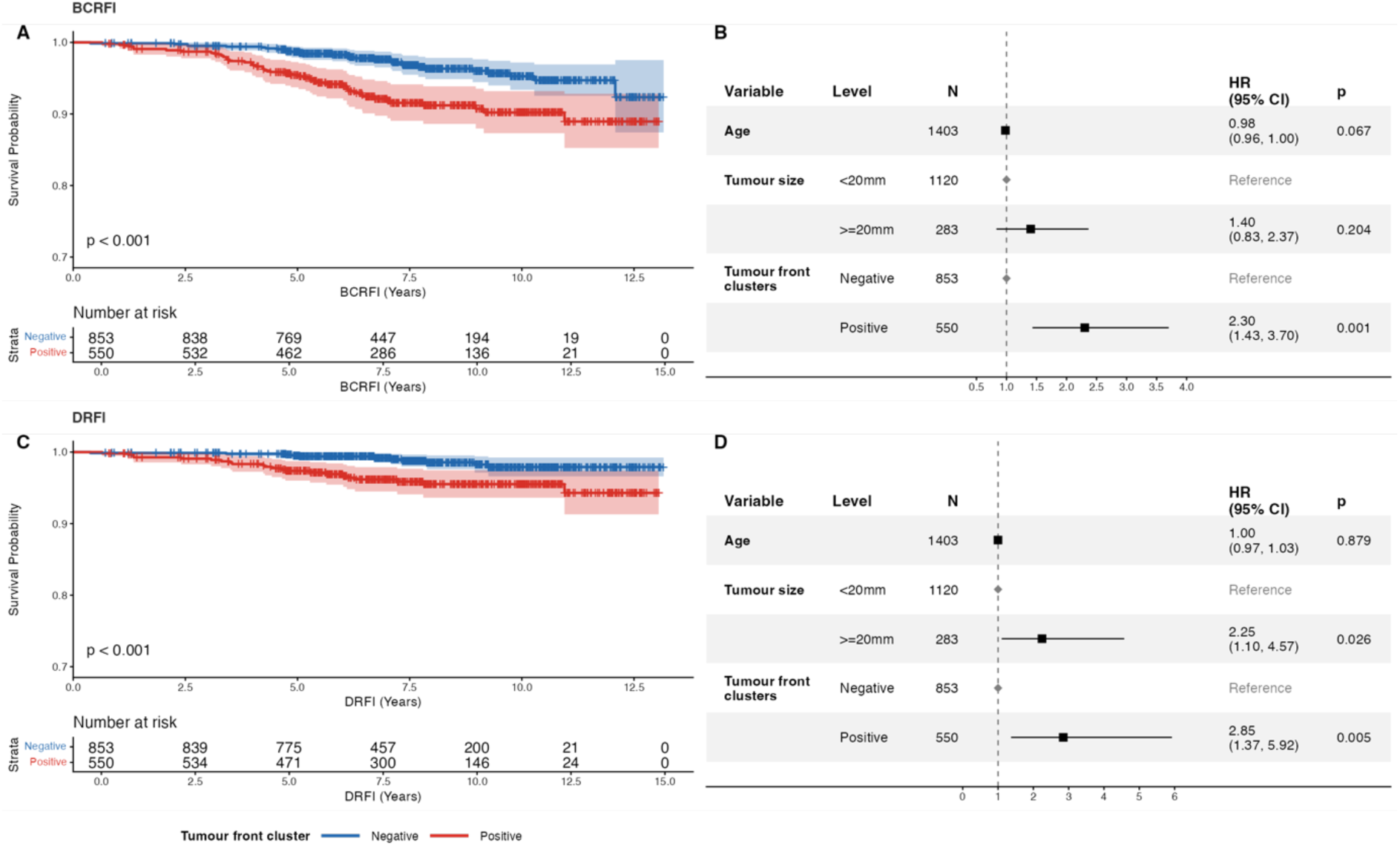
Recurrence-free interval outcomes for breast cancer patients in the external test cohort, ER+/HER2-/LN-/NHG 1&2 subgroup, stratified by tumour front cluster status. Kaplan–Meier curves for breast cancer recurrence-free interval in the independent test set with BCRFI as outcome (A) and DRFI as outcome (C). Forest plots from Cox proportional hazard regression analyses for BCRFI (B) and DRFI (D), adjusted for age and tumour size. BCRFI, Breast Cancer Recurrence-Free Interval; DRFI, Distant metastases Recurrence-Free interval HR, hazard ratio; CI, confidence interval

**Table 1.**
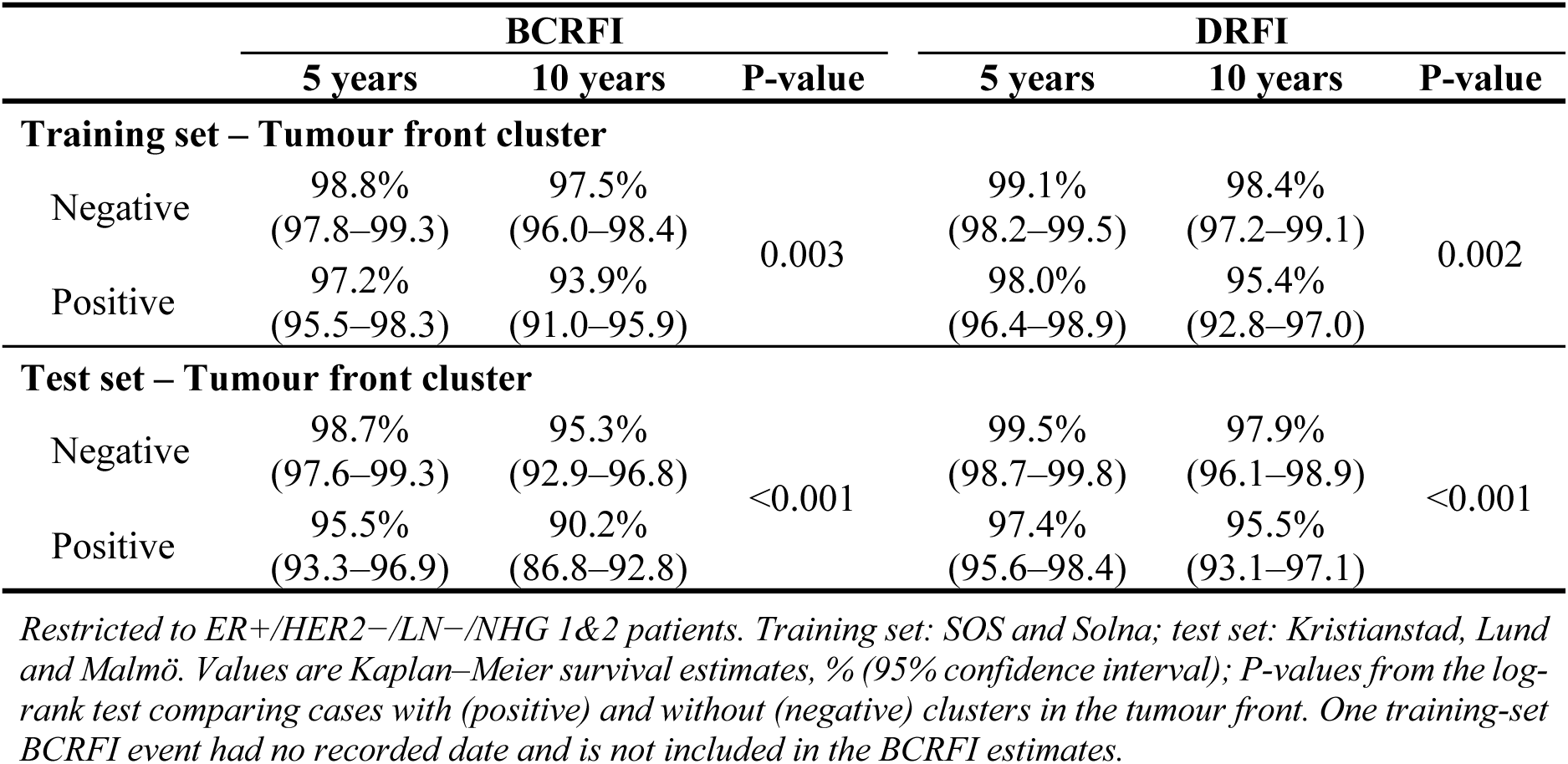
Absolute differences in survival between subgroups in both breast cancer recurrence-free interval (BCRFI) and distant-metastases recurrence-free interval (DRFI) in the train and test cohorts.

### Prognostic value of tumour front clusters in the ViT_DG-low subgroup

To evaluate whether tumour front cluster positivity provides prognostic value beyond overall DeepGrade biomarker status, analyses were repeated in the ER+/HER2-/LN-subgroup classified as ViT_DG-low. In the training cohort (n=1133), cluster positive status was associated with significantly worse BCRFI in univariable analysis (HR 2.61, 95% CI 1.15– 5.93, p = 0.022) with an absolute difference in survival of 2.1% and 3.8% at 5 and 10 years respectively (Supplementary Table S3). However, the multivariable association did not reach statistical significance (HR 2.20, 95% CI 0.96–5.05, p = 0.063). In the external test cohort (n=1041) cluster-positive status showed a strong and statistically significant association with worse BCRFI both in univariable (HR 3.24, 95% CI 1.81–5.81, p < 0.001) and multivariable analysis (HR 2.86, 95% CI 1.56–5.24, p = 0.001). The absolute difference between the cluster-positive and cluster negative groups were of 5.1% and 8.3% at 5 and 10 years respectively. The association with distant-metastases recurrence-free interval was also confirmed in the validation cohort (HR 4.76, 95% CI 1.98–11.44, p < 0.001 univariable; HR 3.57, 95% CI 1.43– 8.90, p = 0.006 multivariable; Supplementary Figure S4). These results suggest that the spatial clustering of high-risk tiles at the tumour front carries prognostic information that is not captured by the global AI-biomarker.

### Benchmarking compared to existing clinical risk groups

To contextualise the absolute prognostic impact of the tumour front cluster marker, the 10-year BCRFI and DRFI in the positive and negative groups were compared to the survival rates in standard clinical risk groups in the same cohorts (Figure 4). In the test cohort cluster-negative patients achieved a 10-year BCRFI of 95.3%, identical to those in the NHG 1 subgroup, and much higher than those in the NHG 2 subgroup (92.4%). Patients in the NHG 2 and cluster negative subgroup have a BCRFI of 94%, therefore above the NHG 2 reference, but lower than NHG 1, showing a reclassification of patients with lower risk. Cluster-positive patients on the other hand showed BCRFI rates of 90.2%, which is lower than the rate in the NHG 3 subgroup (92.6%), and approaches recurrence risk of lymph-node positive patients. This demonstrates that tumour front cluster status could reclassify patients into clinically meaningful subgroups within the NHG1-2 category. In the full unselected cohorts, cluster-positive status remained associated with BCRFI after adjustment for age, tumour size, NHG, lymph node status, ER and HER2 status (test HR 1.49, 95% CI 1.06–2.09), whereas the association with DRFI was significant in the training but not the test cohort (HR 1.46, 95% CI 0.96–2.24; Supplementary Figures S5 and S6).

**Figure 4.**
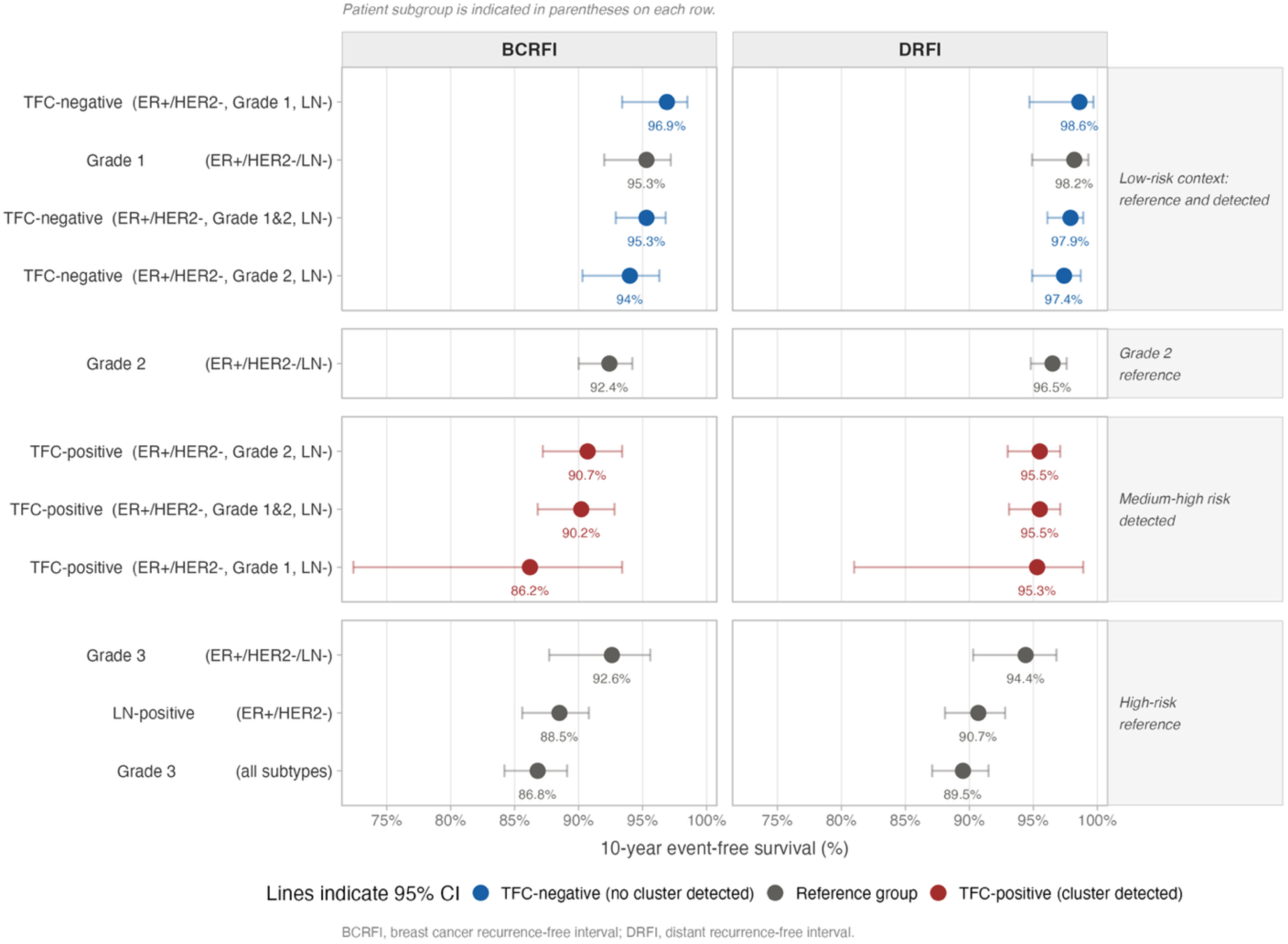
Ten-year breast cancer recurrence-free interval (BCRFI) and distant metastases recurrence-free interval (DRFI) for cluster-stratified (TFC) patients in different subgroups benchmarked against established clinical risk groups, in the independent test cohort.

## Discussion

In this study we developed and validated a novel AI-based method to explicitly model focal clusters of a previously reported prognostic AI-based biomarker, demonstrating that the spatial distribution of high-grade morphology at the invasive tumour front carries independent prognostic information. In the primary subgroup of ER+/HER2-, NHG1 and 2, lymph node negative patients, tumour front cluster status stratified patients into groups with significant differences in recurrence rates. This is the subgroup of patients in which treatment decisions are the hardest, and in which the risk of late recurrence is poorly predicted by the standard clinicopathological factors, and in particular grade^32–34^. This H&E-based spatial biomarker derived from routinely available slides could directly address this unmet need. The finding that tumour front clusters remain prognostic in patients already classified as low risk by ViT_DG is of particular note. In the ER+/HER2-/LN-subgroup classified as ViT_DG-low, cluster positivity was significantly associated with worse survival in the external validation cohort. This indicates that a patient that has a globally low-grade tumour profile can still have some focal regions of high-grade morphological clustering at the invasive front that is associated with a higher recurrence risk. On the other hand, patients with cluster negative status in the ViT_DG-low group represent a subgroup with a very low risk of recurrence. This step-wise stratification, first by the global level AI-biomarker (or a bulk-level molecular assay) and then by spatial clustering, could in principle guide more personalised treatment decisions, identifying patients who may benefit from therapy despite a favourable overall profile, and those in whom de-escalation should be initiated with high confidence.

The clinical and pathological characteristics of cluster-positive patients show biological understandings for the spatial marker. Within the primary subgroup, cluster positive patients showed a higher proportion of ViT_DG-high tumours, higher median Ki67 expression, a higher proportion of NHG2 tumours and larger tumours. While these differences show some biological basis to the spatial marker on a global level, they were modest and the cluster marker remained independent in the multivariable analyses indicating it captures prognostic risk not fully covered by standard clinical variables alone. Co-localised high-grade morphology at the tumour front could show some locally aggressive tumour in the invasive tumour front. Indeed, the tumour front is biologically distinct from the tumour centre and manually assessed tumour budding in that area has been shown to be prognostic^23,24^. That the spatial clustering of the AI-predicted high-grade morphology is specifically enriched in this anatomical zone, and that its clustering is independently prognostic is consistent with the hypothesis that the tumour front could have a subpopulation of cells with disproportionate metastatic potential.

The growing body of evidence that deep-learning predicted intratumour heterogeneity in breast cancer prognosis and the use of Graph Neural Networks^35,36^ supports our approach. Our contribution is to demonstrate that the spatial distribution of a previously validated marker, rather than a newly trained representation, independently adds prognostic value. A similar principle has been investigated for TILs, where it is not only the overall density that is associated with survival, but also their localisation and spatial clustering^37^. Our study shows that this clustering effect on survival prediction is also true for AI-derived morphological markers, emphasising that not only the overall level of a tumour biomarker matters, but also its spatial localisation in the tumour.

This is one of the largest retrospective H&E-based prognostic studies in early breast cancer with over 3000 patients in both the training and test sets, from five different hospitals with a fully independent test set drawn from a prospective population-based study from a geographically and administratively different region. It builds upon an existing biomarker, DeepGrade, adapted to the most recent state of the ViT-based feature extraction. There are also some limitations. The analysis is retrospective, and although the test cohort was drawn from a prospective, population-based study with the tumour front cluster marker was locked before validation, the clinical variables depend on registry data, and on slides being retrieved from routine archives. Adjuvant treatment was neither randomised nor standardised, so treatment received may have modified recurrence risk. Given the high disproportinality in breast cancer prevalence between genders, the study only include female patients. Finally, given the nature of the analyses and the spatial area required, some very small tumours might have been excluded, potentially leading to a larger effect size if they had been included as these tumours are likely to be of the lower risk.

## Conclusion

The results obtained demonstrate that the presence of clusters of high-risk tiles at the invasive tumour front constitutes an independent prognostic factor for breast cancer recurrence-free interval and distant-metastasis recurrence-free interval in ER+/HER2−, LN−, NHG 1–2 breast cancer patients, in both the training and independent test cohorts. This suggests that the spatial localisation of high-risk morphological patterns and specifically its high concentration at the tumour front, captures prognostic information beyond the overall proportion of high risk tiles. These findings motivate the hypothesis that spatially localised signals may be attenuated when a tumour is summarised as a single average value, as in bulk molecular profiling of, for example, tumour mRNA expression, or with a single AI-based score. The spatial cluster marker enables identification of a high-risk subset within patients otherwise classified as low-to-intermediate risk, the clinical subgroup where decision making is currently the most uncertain and where the potential of additional risk stratification through affordable biomarkers is of greatest interest. On the other hand, cluster-negative patients achieve outcomes in line with the best-prognosis grade 1 group, supporting de-escalation. These findings suggest that spatially-aware AI biomarkers have the potential to be used as a complement for current risk profiling in breast cancer.

## Data Availability

The data are not publicly available, but are available from the corresponding author on reasonable request, subject to ethical approval.

## Acknowledgements

The authors would like to acknowledge patients and clinicians participating in the SCAN-B study, personnel at the central SCAN-B laboratory at the Division of Oncology, Lund University, the Swedish National Breast Cancer Quality Registry (NKBC), Regional Cancer Center South, and the South Sweden Breast Cancer Group (SSBCG). We would also like to acknowledge that this work was conducted as part of a collaborative postdoctoral fellowship between AstraZeneca and Karolinska Institutet.

## Supplementary Figures and Tables

**Supplementary Figure S1.**
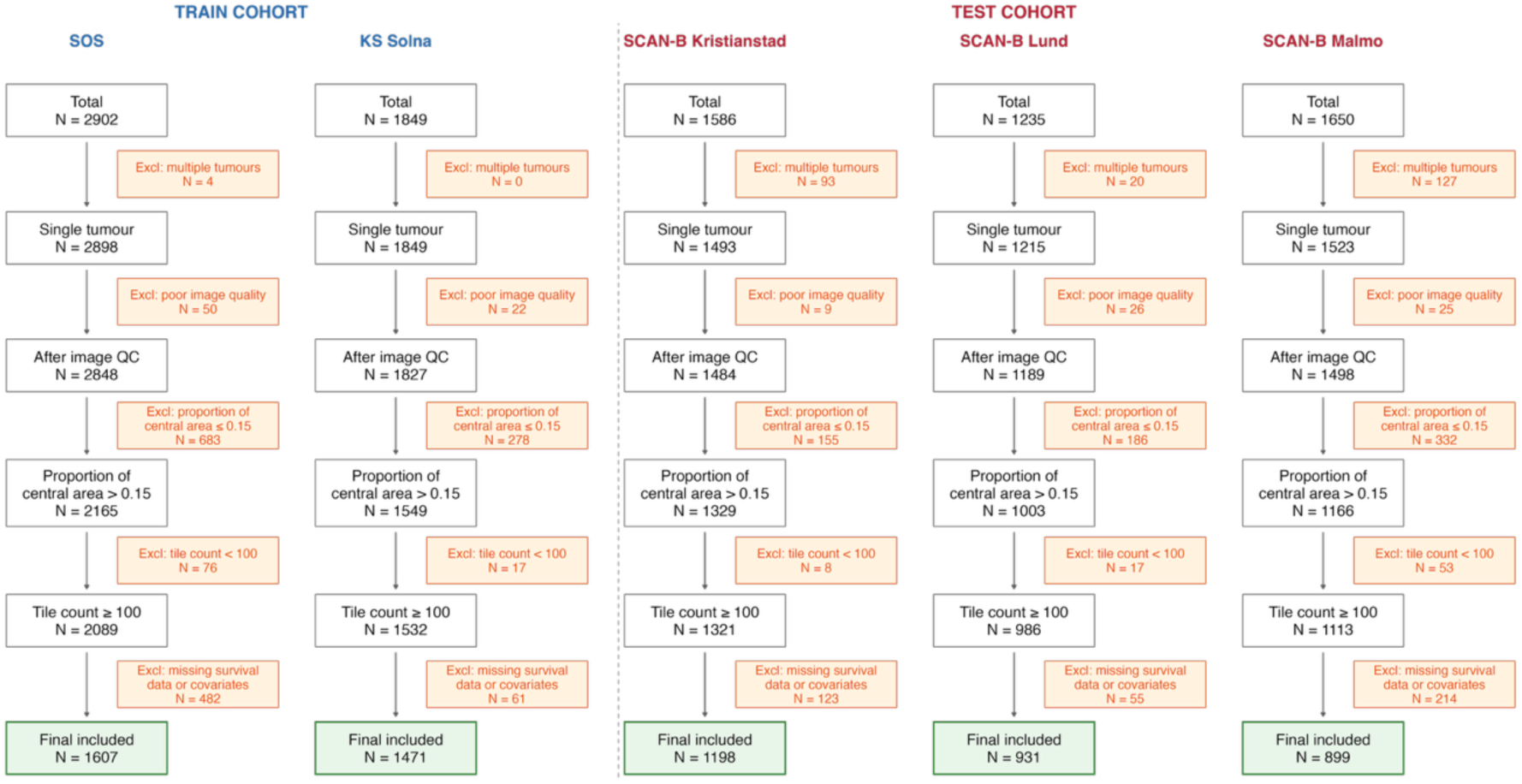
Flowchart illustrating the process of patient selection for each clinical site individually.

**Supplementary Table S1.** Clinical characteristics for patients in each cohort.

| Characteristic | Train<br>SOS | Solna | Test<br>Kristianstad | Lund | Malmo |
| --- | --- | --- | --- | --- | --- |
| N | 1607 | 1471 | 1198 | 931 | 899 |
| *Age* | 61.75 ± 11.73 | 61.14 ± 12.28 | 66.62 ± 12.42 | 65.12 ± 12.55 | 65.19 ± 13.05 |
| *Histologic grade* |  |  |  |  |  |
| NHG1 | 348 (21.66) | 319 (21.69) | 208 (17.36) | 183 (19.66) | 169 (18.80) |
| NHG2 | 772 (48.04) | 753 (51.19) | 641 (53.51) | 459 (49.30) | 492 (54.73) |
| NHG3 | 487 (30.30) | 399 (27.12) | 349 (29.13) | 289 (31.04) | 238 (26.47) |
| *Tumour size* |  |  |  |  |  |
| <20mm | 1037 (64.53) | 967 (65.74) | 760 (63.44) | 639 (68.64) | 589 (65.52) |
| >=20mm | 570 (35.47) | 504 (34.26) | 438 (36.56) | 292 (31.36) | 310 (34.48) |
| *Lymph node* |  |  |  |  |  |
| Negative | 1165 (72.50) | 1095 (74.44) | 815 (68.03) | 660 (70.89) | 598 (66.52) |
| Positive | 442 (27.50) | 376 (25.56) | 383 (31.97) | 271 (29.11) | 301 (33.48) |
| *ER status* |  |  |  |  |  |
| Negative | 143 (8.90) | 175 (11.90) | 153 (12.77) | 100 (10.74) | 104 (11.57) |
| Positive | 1464 (91.10) | 1296 (88.10) | 1045 (87.23) | 831 (89.26) | 795 (88.43) |
| *HER2 status* |  |  |  |  |  |
| Negative | 1428 (88.86) | 1317 (89.53) | 1067 (89.07) | 848 (91.08) | 779 (86.65) |
| Positive | 176 (10.95) | 152 (10.33) | 128 (10.68) | 83 (8.92) | 108 (12.01) |
| NA | 3 (0.19) | 2 (0.14) | 3 (0.25) | 0 (0.00) | 12 (1.33) |
| *KI67 status* |  |  |  |  |  |
| Low | 759 (47.23) | 462 (31.41) | 552 (46.08) | 397 (42.64) | 255 (28.36) |
| High | 763 (47.48) | 472 (32.09) | 353 (29.47) | 351 (37.70) | 527 (58.62) |
| NA | 85 (5.29) | 537 (36.51) | 293 (24.46) | 183 (19.66) | 117 (13.01) |
| *ViT_DG* |  |  |  |  |  |
| Low | 717 (44.62) | 791 (53.77) | 571 (47.66) | 476 (51.13) | 449 (49.94) |
| High | 890 (55.38) | 680 (46.23) | 627 (52.34) | 455 (48.87) | 450 (50.06) |
| *Tumour front cluster* |  |  |  |  |  |
| Negative | 623 (38.77) | 686 (46.63) | 447 (37.31) | 392 (42.11) | 377 (41.94) |
| Positive | 984 (61.23) | 785 (53.37) | 751 (62.69) | 539 (57.89) | 522 (58.06) |
| *BCRFI events* |  |  |  |  |  |
| No | 1508 (93.84) | 1359 (92.39) | 1083 (90.40) | 870 (93.45) | 828 (92.10) |
| Yes | 99 (6.16) | 112 (7.61) | 115 (9.60) | 61 (6.55) | 71 (7.90) |
| *DRFI events* |  |  |  |  |  |
| No | 1521 (94.65) | 1380 (93.81) | 1120 (93.49) | 885 (95.06) | 848 (94.33) |
| Yes | 86 (5.35) | 91 (6.19) | 78 (6.51) | 46 (4.94) | 51 (5.67) |

*NHG* Nottingham histological grade, *ER* Estrogen receptor, *HER2* human epidermal growth factor 2, *KI67* Ki-67 proliferation index (Low <20%, High ≥20%), *ViT\_DG* vision transformer DeepGrade, *BCRFI* breast-cancer recurrence-free interval, *DRFI* distant-metastases recurrence-free interval, *NA* not available.

**Supplementary Table S2.** Clinical characteristics by train/test split for the primary subgroup analyses.

| Characteristic | Overall | Test | Train |
| --- | --- | --- | --- |
| N | 2939 | 1403 | 1536 |
| **Age** | 64.36 ± 11.41 | 66.46 ± 11.65 | 62.45 ± 10.85 |
| **Tumour size** |  |  |  |
| <20mm | 2294 (78.05) | 1120 (79.83) | 1174 (76.43) |
| ≥20mm | 645 (21.95) | 283 (20.17) | 362 (23.57) |
| **KI67 status** |  |  |  |
| Low | 1727 (58.76) | 821 (58.52) | 906 (58.98) |
| High | 693 (23.58) | 359 (25.59) | 334 (21.74) |
| NA | 519 (17.66) | 223 (15.89) | 296 (19.27) |
| **ViT_DG** |  |  |  |
| Low | 2096 (71.32) | 1004 (71.56) | 1092 (71.09) |
| High | 843 (28.68) | 399 (28.44) | 444 (28.91) |
| **Tumour front cluster** |  |  |  |
| Negative | 1825 (62.10) | 853 (60.80) | 972 (63.28) |
| Positive | 1114 (37.90) | 550 (39.20) | 564 (36.72) |
| **BCRFI events** |  |  |  |
| No | 2817 (95.85) | 1330 (94.80) | 1487 (96.81) |
| Yes | 122 (4.15) | 73 (5.20) | 49 (3.19) |
| **DRFI events** |  |  |  |
| No | 2871 (97.69) | 1370 (97.65) | 1501 (97.72) |
| Yes | 68 (2.31) | 33 (2.35) | 35 (2.28) |

Restricted to ER+/HER2-/LN-/NHG 1&2 patients. Train set: SOS and Solna sites. Test set: all remaining sites. KI67 Ki-67 proliferation index (Low <20%, High ≥20%), ViT\_DG vision transformer DeepGrade, BCRFI breast-cancer recurrence-free interval, DRFI distant-metastases recurrence-free interval, NA not available.

**Supplementary Figure S2.**
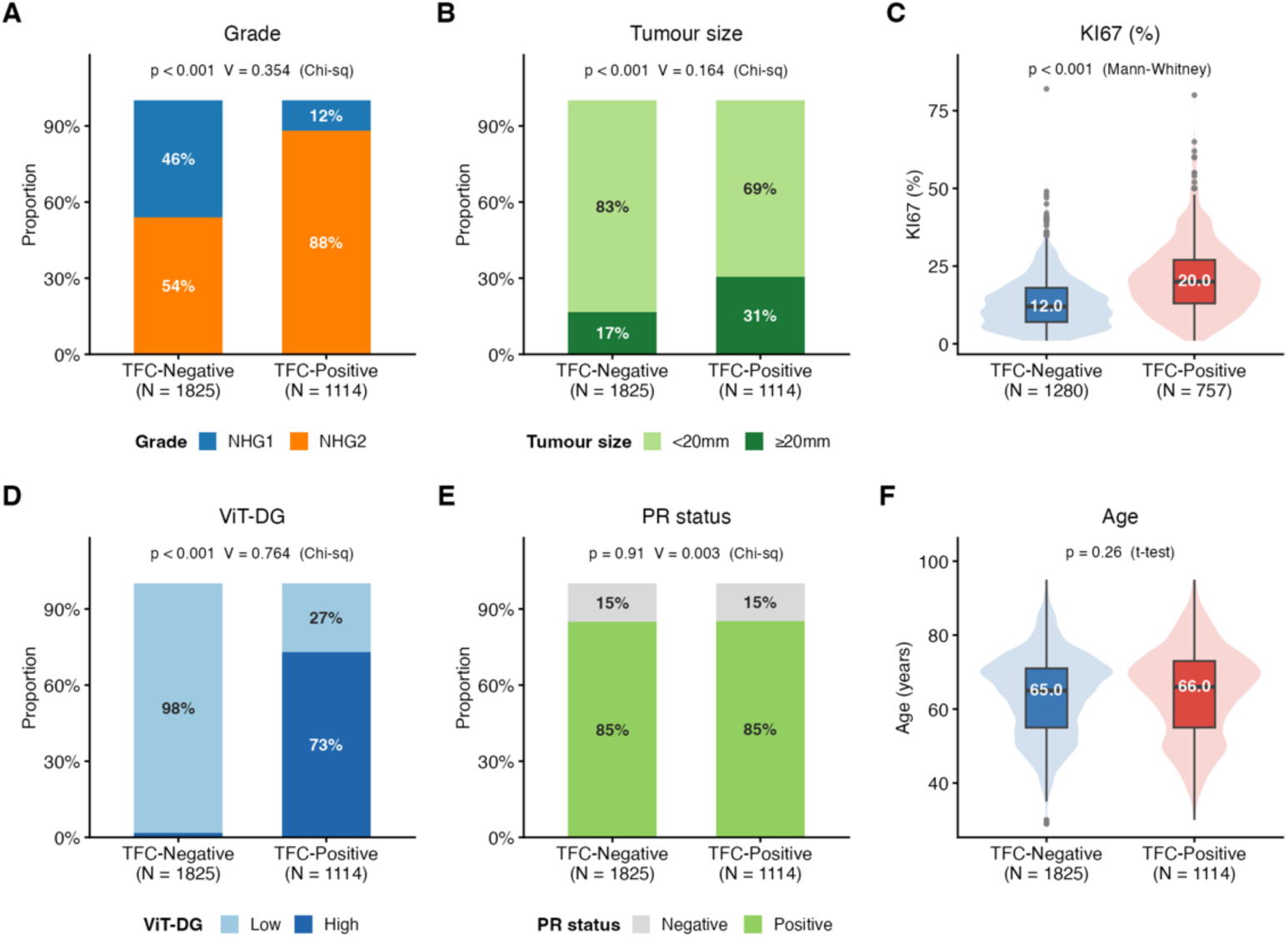
Clinical and pathological characterisation of Tumour front cluster-positive and Tumour front cluster-negative patients in the ER+/HER2−/LN−/Grade 1&2 subgroup (whole cohort; N =2939). Bar charts show distribution of NHG, ViT_DG status, PR status, KI67 status, ltumour size and age. Violin plots show age and KI67 percentage. P-values are from Fisher’s exact test, chi-squared test, t-test, or Mann–Whitney U test as indicated. V, Cramér’s V effect size.

**Supplementary Table S3.** Absolute differences in survival between subgroups in both breast cancer recurrence-free interval (BCRFI) and distant-metastases recurrence-free interval (DRFI) in the ER+/HER2−/LN−/ViT_DG-low subgroup in the train and test cohorts.

|  | BCRFI |  |  | DRFI |  |  |
| --- | --- | --- | --- | --- | --- | --- |
|  | 5 years | 10 years | P-value | 5 years | 10 years | P-value |
| Training set – Tumour front cluster |  |  |  |  |  |  |
| Negative | 98.8% | 97.6% | 0.017 | 99.1% | 98.4% | 0.096 |
|  | (97.8–99.3) | (96.2–98.5) |  | (98.2–99.5) | (97.2–99.1) |  |
| Positive | 96.7% | 93.8% |  | 98.7% | 95.8% |  |
|  | (92.3–98.6) | (87.5–97.0) |  | (94.9–99.7) | (89.5–98.3) |  |
| Test set – Tumour front cluster |  |  |  |  |  |  |
| Negative | 98.7% | 95.6% | <0.001 | 99.5% | 98.3% | <0.001 |
|  | (97.6–99.3) | (93.4–97.1) |  | (98.7–99.8) | (96.6–99.1) |  |
| Positive | 93.6% | 87.3% |  | 95.9% | 93.6% |  |
|  | (88.7–96.4) | (80.6–91.9) |  | (91.6–98.0) | (88.3–96.6) |  |

*Restricted to ER+/HER2-/LN-/ViT DG-low patients. Training set: SOS and Solna; test set: Kristianstad, Lund and Malmö. Values are Kaplan–Meier survival estimates, % (95% confidence interval); P-values from the log-rank test comparing cases with (positive) and without (negative) clusters in the tumour front. One training-set BCRFI event had no recorded date and is not included in the BCRFI estimates.*

**Supplementary Figure S3.**
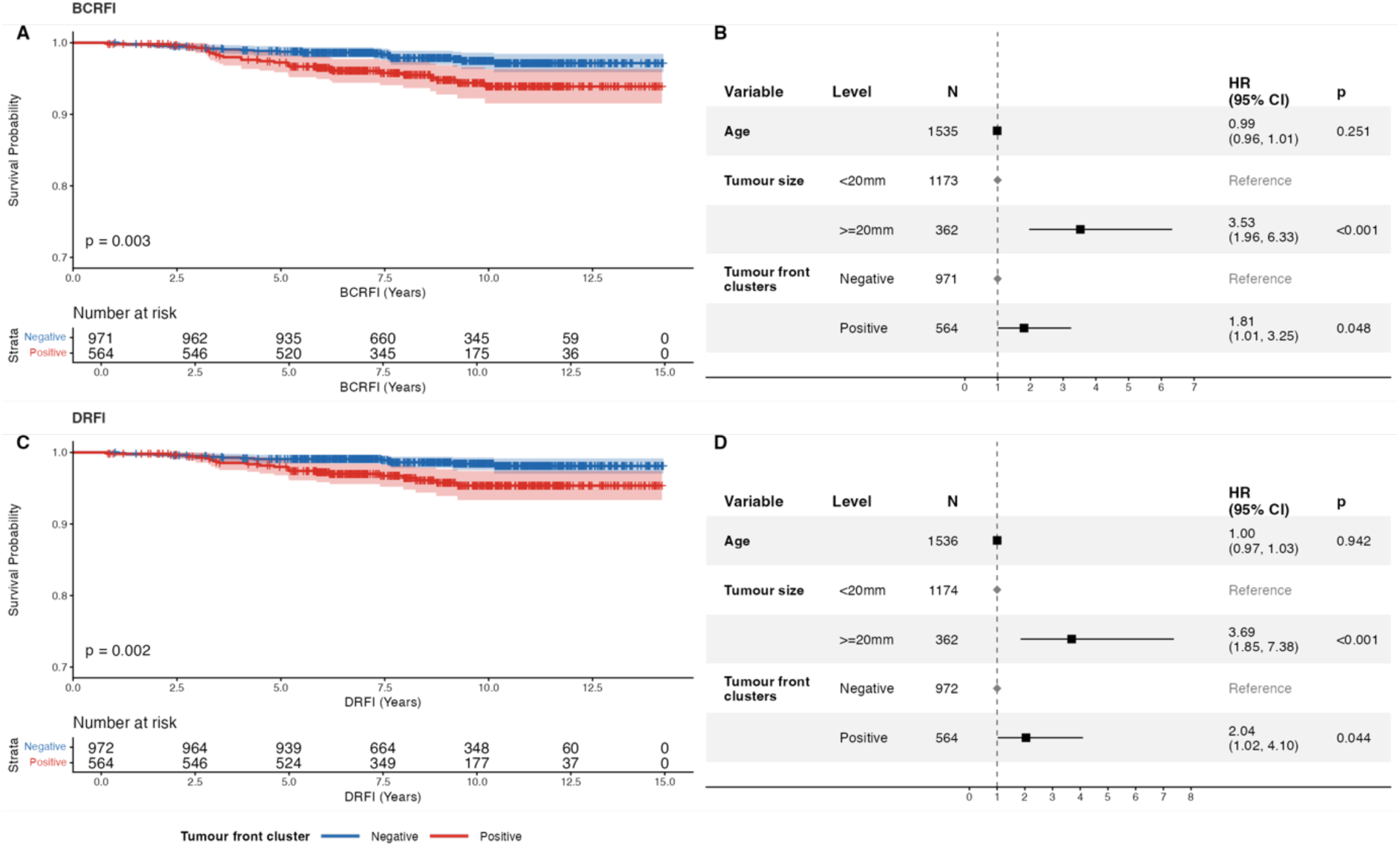
Recurrence-free interval outcomes for breast cancer patients in the ER+/HER2-/LN-/NHG 1&2 subgroup of the training cohort, stratified by tumour front cluster status. Kaplan–Meier curves for breast cancer recurrence-free interval in the training set with BCRFI as outcome (A) and DRFI as outcome (C). Forest plots from Cox proportional hazard regression analyses for BCRFI (B) and DRFI (D), adjusted for age and tumour size. BCRFI, Breast Cancer Recurrence-Free Interval; DRFI, Distant metastases Recurrence-Free interval HR, hazard ratio; CI, confidence interval

**Supplementary Figure S4.**
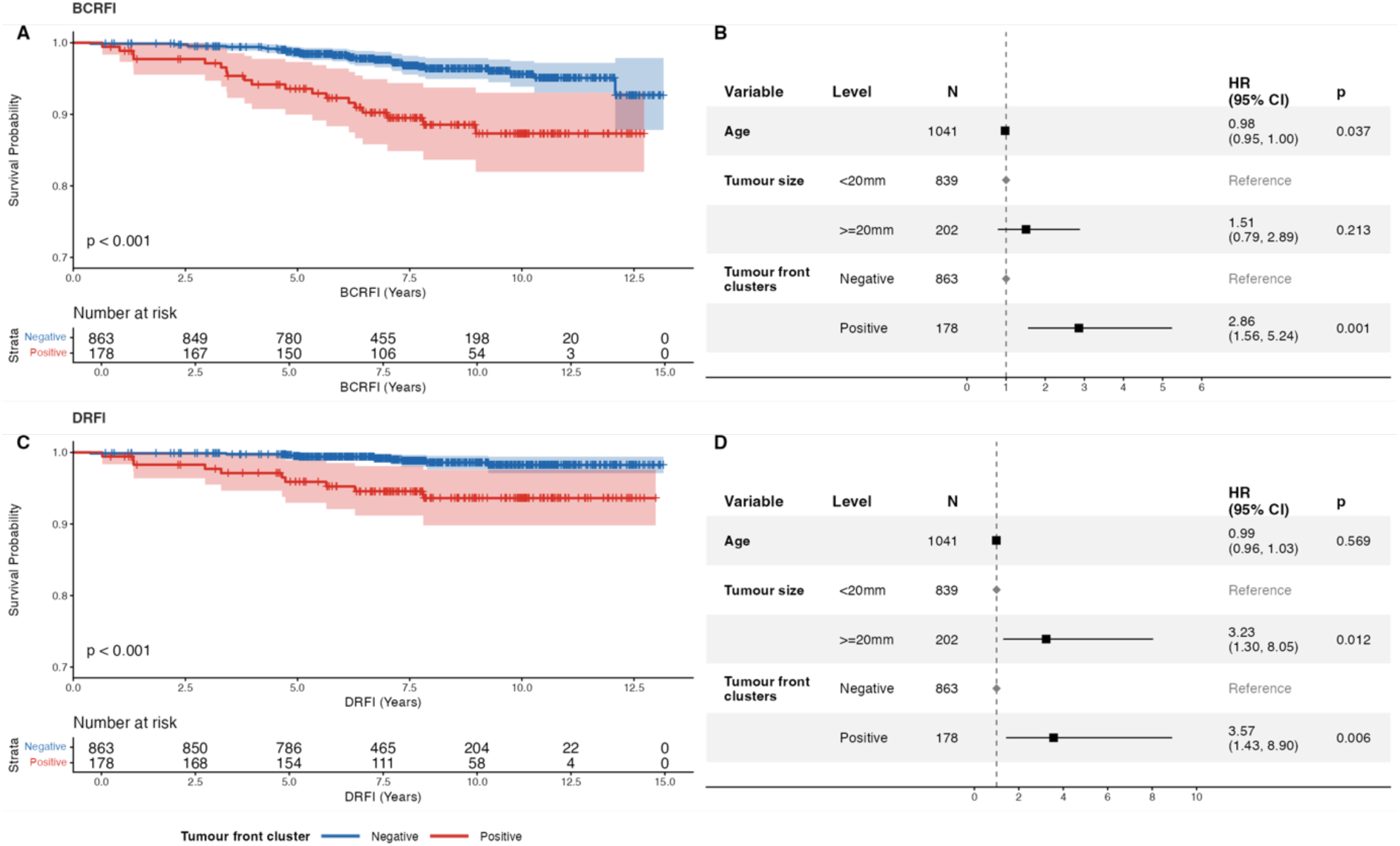
Recurrence-free survival outcomes for breast cancer patients in the external independent testing cohort ER+/HER2-/LN-/ViT_DG-low subgroup, stratified by tumour front cluster status. Kaplan–Meier curves for breast cancer recurrence-free interval (A) and distant-metastasis recurrence-free interval (C). Forest plots from Cox proportional hazard regression analyses for BCRFI (B) and for DRFI (D), adjusted for age and tumour size. HR, hazard ratio; CI, confidence interval

**Supplementary Figure S5.**
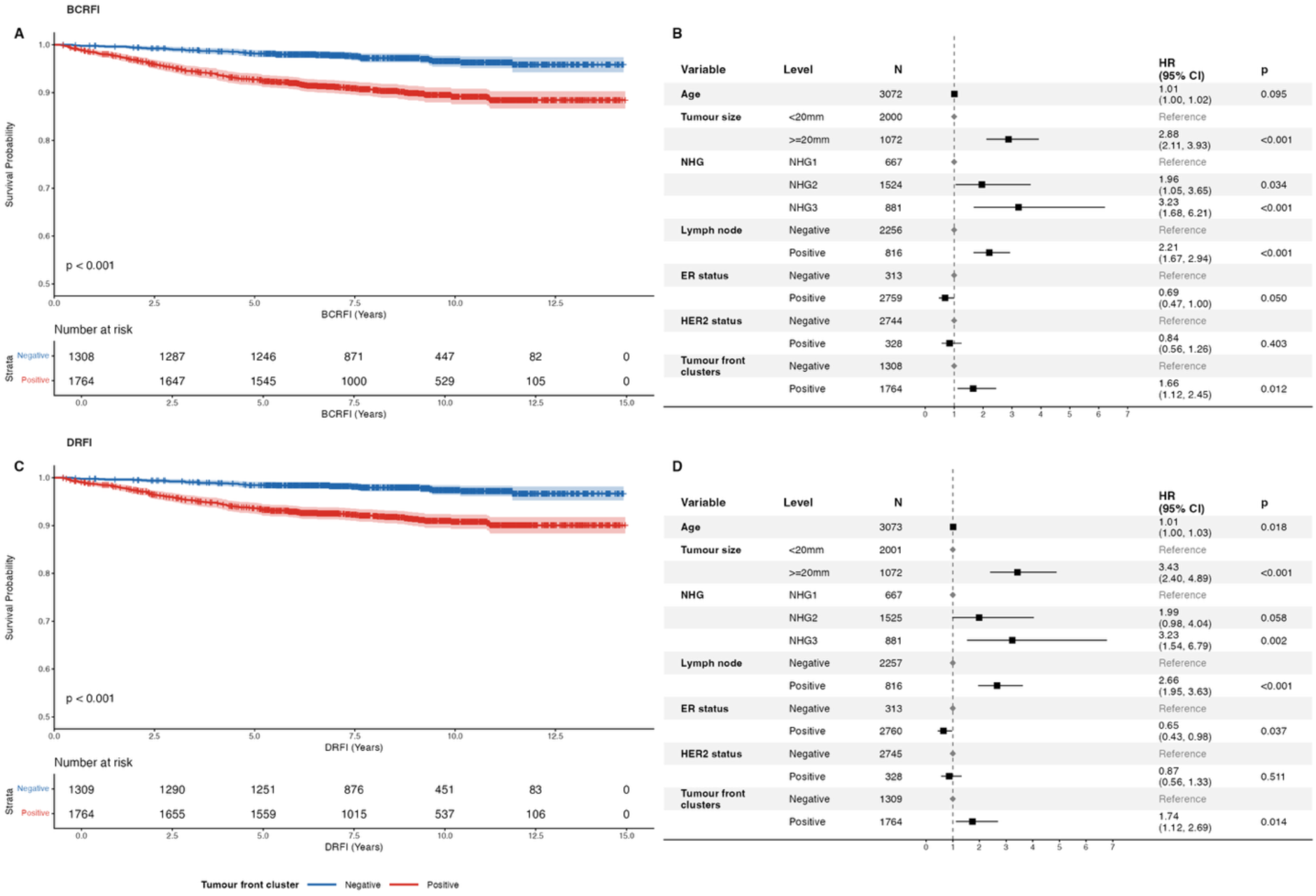
Recurrence-free survival outcomes for breast cancer patients in the full training cohort, stratified by tumour front cluster status. Kaplan–Meier curves for breast cancer recurrence-free interval (A) and distant-metastasis recurrence-free interval (C). Forest plots from Cox proportional hazard regression analyses for BCRFI (B) and for DRFI (D), adjusted for age and tumour size. HR, hazard ratio; CI, confidence interval

**Supplementary Figure S6.**
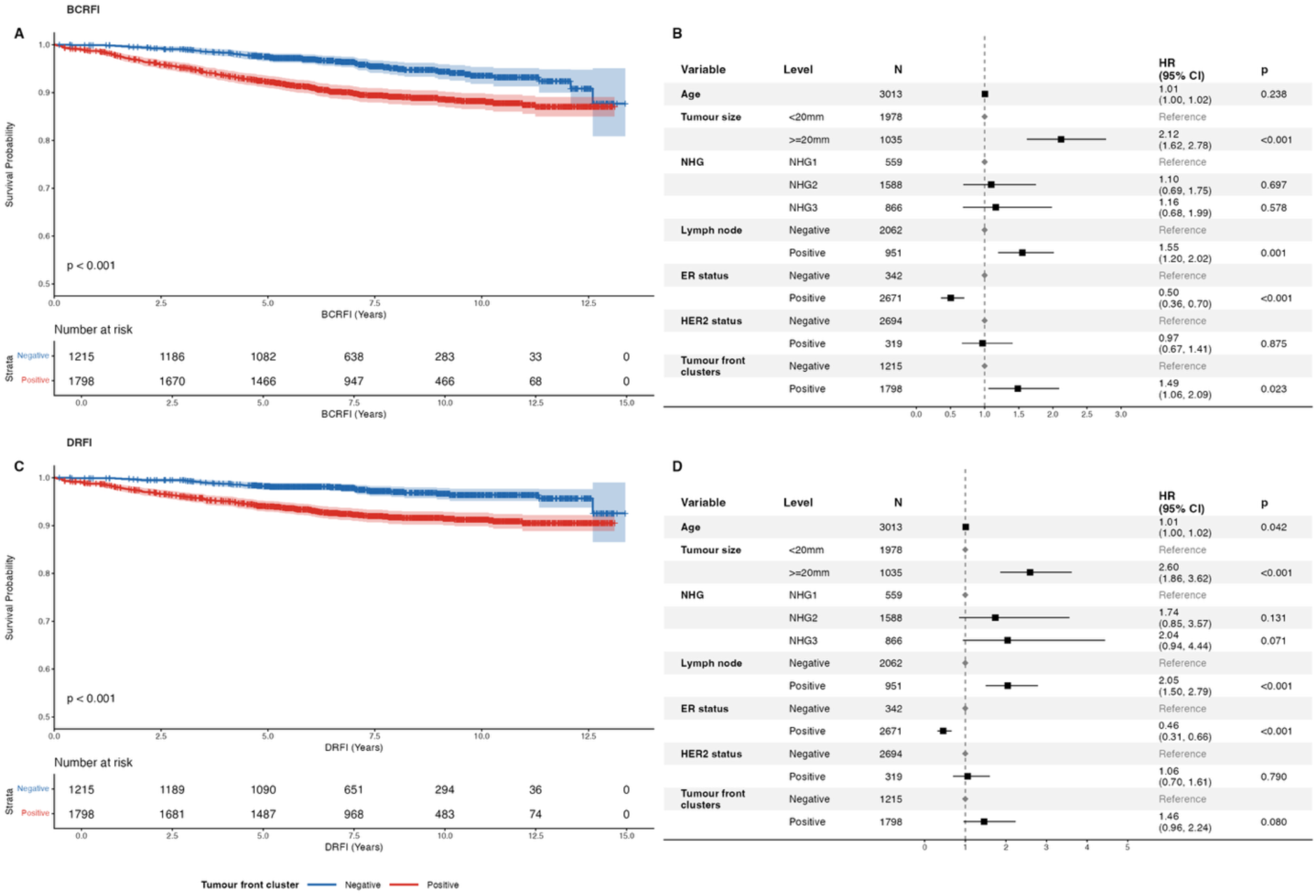
Recurrence-free survival outcomes for breast cancer patients in the full external test cohort, stratified by tumour front cluster status. Kaplan–Meier curves for breast cancer recurrence-free interval (A) and distant-metastasis recurrence-free interval (C). Forest plots from Cox proportional hazard regression analyses for BCRFI (B) and for DRFI (D), adjusted for age and tumour size. HR, hazard ratio; CI, confidence interval

